# The Impact of Freeze-Thaw Cycles on the Integrity of SARS-COV-2 Viral Culture Fluids and Clinical Remnant Samples in Antigen or Nucleic Acid Testing

**DOI:** 10.1101/2022.12.14.22282041

**Authors:** Hajirah Noor Hussain, Hali Weeks, Derek Zhou, Divya Joseph, Brooke Lam, Haidong Xu, Chushi Zhang, Keqin Gregg, Wenli Zhou

**Affiliations:** XYZ Laboratory, 2009 Ranch Rd. 620 N, STE 325, Lakeway, TX 78734

**Keywords:** Freeze-thaw cycle, SARS-CoV-2, RT-PCR, lateral flow assay, rapid antigen test

## Abstract

**Preservation at ultra-low temperatures has been a gold standard** for long-term storage of many types of clinical specimens including the SARS-CoV-2 virus. The frozen specimens can be easily transported and tested later. In addition, de-identified frozen remnant samples are resources for many preclinical or clinical studies. It is therefore crucial to understand whether freeze and thaw cycles (FTCs) can adversely affect SARS-CoV-2 test performance when frozen samples are tested. Some early studies suggest that the FTCs increased the cycles threshold (Ct) of RT-PCR indicating the potential degradation of the SARS-CoV-2 nucleic acid after FTCs, while the others did not report any significant changes in the SARS-CoV-2 nucleic acids after the FTCs. Moreover, the impact of FTCs on the performance of the SARS-CoV-2 antigen test is scarcely reported.

**In this study, we performed paired nucleic acid and rapid antigen tests** on the same samples to investigate and directly compare how FTCs affect the performance of two types of tests. Both inactivated viral culture fluid samples and clinical remnant samples were studied. Our results showed that FTCs had minimal negative effects on the performance of the rapid SARS-CoV-2 antigen test, and the test results remained largely consistent throughout the FTCs, whereas the Ct values of RT-PCR increased with the increase of the FTC numbers. In addition, our data also demonstrated that the SARS-CoV-2 is preserved better in VTM than PBS during FTCs in regard to nucleic acid testing.

## Introduction

**Since the COVID-19 pandemic began** two and half years ago, a large number of studies have been conducted on SARS-CoV-2 due to its high infectivity, mutability, and the need for faster and more accurate methods of testing. The main SARS-CoV-2 testing technologies include reverse transcription real-time PCR (RT-PCR or RT-qPCR), which is the gold standard and tests for viral nucleic acid usually in clinical laboratories, and lateral flow rapid assays (LFA) testing for viral antigens, most often used in point-of-care or home settings. There are times that frozen patient samples must be used for various reasons such as delayed testing, secured transportation, late verification, or research and development using remnant samples, freezing at an ultra-low temperature remains the only option for long-term specimen storage as per CDC recommendation (https://www.cdc.gov/coronavirus/2019-nCoV/lab/guidelines-clinical-specimens.html). It is thus crucial to fully understand whether FTCs can adversely affect the test performance on any scale if frozen samples are used.

**Several studies using clinical samples** (1-2), wastewater solid (3-4), and synthetic RNA (5) showed that FTCs had a significant, negative impact on the Ct values of the SARS-CoV-2 RT-PCR test, indicating a degradation of the viral nucleic acids throughout FTCs even with only one freeze and thaw cycle. Others found that a limited number of FTCs had a minor or no significant impact on the RT-PCR test results (6-9). There are fewer studies of the FTC’s impact on the lateral flow rapid antigen tests. Cubas-Atienzar et al. (10) reported that 11 out of 19 rapid antigen tests examined had at least a 2-fold decrease in sensitivity even after just one FTC and 2 of 19 showed increased sensitivity. Zhou et al. (11) found a slight decrease in antigen test signal intensity after 2-3 FTCs in three rapid antigen tests used in that study. Both studies indicated that the types of rapid test kits and diluent used in the studies could have a significant impact on the test performance after the FTCs.

**There are times that VTM preserved SARS-CoV-2 samples** need to be tested by both RT-PCR and rapid antigen methods for test result confirmation or during assay development. It becomes important to understand how FTCs would differently affect the RT-PCR and antigen tests under the same sample conditions. In this study, we seek to answer this question by testing clinical remnant samples and inactivated viral fluid of various strains using the RT-PCR method as well on a rapid antigen test kit. Our approach and conclusion shall provide a reference for using frozen samples on different tests as well as a model for any future RT-PCR and antigen comparison studies of FTC’s impact.

## Materials and Methods

### Overall Study Design

**The study was performed with two types of specimens**, one was various strains of inactivated viral culture fluids, and another was clinical remnant samples. For inactivated viral culture fluids, the limit of detection (LoD) was first determined using the rapid antigen kit (Hotgen Coronavirus (2019-nCoV)-Antigen test- (Beijing Hotgen Biotech Co. Ltd., Beijing, China), or Antigentest) for each viral culture sample. Contrived samples at various low to middle concentrations along with a negative matrix were aliquoted, then subjected to various FTCs. For the clinical remnant samples, no dilutions were performed. The remnant samples had various Ct values representing high and low viral loads; these were directly aliquoted and subjected to various FTCs. Enough aliquot volume for each sample was made so that each aliquoted sample could be used for both the RT-PCR and antigen tests after the designated number of FTCs.

### Viral Culture Fluid

**Table 1.** listed inactivated SARS-CoV-2 viral culture fluid reference materials used in this study, by variant, catalog #, manufacturer, inactivation method, and stock concentration.

| **Sample ID** | **Variant/Isolate** | **Catalog #** | **Manufacturer** | **Inactivation method** | **Stock Concentration** |
| --- | --- | --- | --- | --- | --- |
| **CS01** | SARS-CoV-2<br>USA-WA1/2020 | 0810587UV | ZeptoMetrix,<br>Buffalo, NY | UV-inactivated | $1.15 \times 10^7$<br>TCID<sub>50</sub>/mL |
| **CS02** | SARS-CoV-2<br>USA-WA1/2020 | 0810587CFHI | ZeptoMetrix,<br>Buffalo, NY | Heat-inactivated | $1.15 \times 10^7$<br>TCID<sub>50</sub>/mL |
| **CS03** | SARS-CoV-2<br>USA-WA1/2020 | 0810587CFHI | ZeptoMetrix,<br>Buffalo, NY | Heat-inactivated | $3.16 \times 10^6$<br>TCID<sub>50</sub>/mL |
| **CS04** | SARS-CoV-2<br>USA-WA1/2020 | 0810587CFHI | ZeptoMetrix,<br>Buffalo, NY | Heat-inactivated | $8.51 \times 10^7$<br>TCID<sub>50</sub>/mL |
| **CS05** | Variant B.1.1.7<br>England/204820<br>464/2020 | 0810614UV | ZeptoMetrix,<br>Buffalo, NY | UV-inactivated | $3.80 \times 10^6$<br>TCID<sub>50</sub>/mL |
| **CS06** | Variant B.1.351<br>South_Africa/K<br>RIS-K005325/2020 | 0810613UV | ZeptoMetrix,<br>Buffalo, NY | UV-inactivated | $1.15 \times 10^7$<br>TCID<sub>50</sub>/mL |
| **CS07** | Variant<br>B.1.617.2, Delta<br>USA/PHC658/20<br>21 | 0810624UV | ZeptoMetrix,<br>Buffalo, NY | UV-inactivated | $1.15 \times 10^7$<br>TCID<sub>50</sub>/mL |
| **CS08** | Variant<br>B.1.1.529<br>Omicron<br>USA/GA-EHC-2811C/2021 | NR-56496 | BEI Resources,<br>Manassas, VA | Gamma-inactivated | $1.51 \times 10^6$<br>TCID<sub>50</sub>/mL |
| **CS09** | Omicron Variant<br>USA/GA-EHC-2811C/2021 | VR-3347 | ATCC,<br>Manassas, VA | Heat-inactivated | $1.9 \times 10^6$<br>genetic copies/uL |

### Negative Matrix

**The negative matrix serves as a diluent for the SARS-CoV-2 culture fluids** and mimics the environment in the human nasal cavity. The nasal swabs from multiple, presumably SARS-CoV-2-negative individuals were collected, eluted in saline (VWR International, Radnor, PA), and pooled. The negative matrix was confirmed by laboratory-developed RT-PCR to be negative.

### Remnant Clinical Samples

**A total of 25 de-identified remnant COVID-19 testing samples**, 20 positives, and 5 negatives were used for the study. These samples were collected using anterior nasal swabs in a PBS or VPM buffer (Lampire, Pipersville, PA). All patients consented and their de-identified remnant samples could be used for the research after testing.

### Freeze-Thaw Cycle Procedure

**The remnant samples were tested initially to establish a baseline**, then aliquoted and placed into corresponding freeze-thaw cycle boxes: 2 FTCs (2c), 4 FTCs (4c), 6 FTCs (6c), 8 FTCs (8c), and 10 FTCs (10c), and stored at -80°C. To perform the FTCs, each box was taken out and thawed to room temperature before they were refrozen, following the scheme in Figure 1.

**Figure 1.**
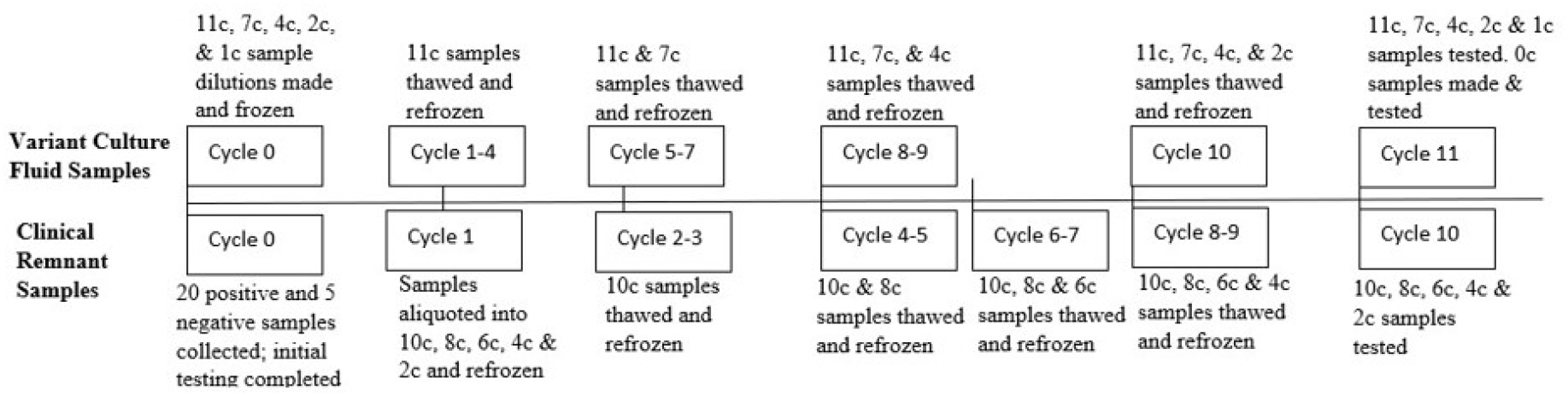
Diagram of FTCs for SARS-CoV-2 Variant and Clinical Remnant Samples. The samples were placed in boxes corresponding to the number of FTCs they underwent before final testing.

**For the inactivated SARS-CoV-2 culture fluid variant sample preparation**, 350 uL of diluted viral cultural fluid with a final concentration of 1.5 to 10 x LoD (on the Antigentest) from each variant strain was aliquoted for a total of 13 samples per cycle and placed in the corresponding freeze-thaw cycle boxes: 1 FTCs (1c), 2 FTCs (2c), 4 FTCs (4c), 7 FTCs (7c), or 11 FTCs (11c) and frozen at -80°C. Each box was taken out and thawed at room temperature before being refrozen to its designated number of FTCs, as shown in Fig. 1.

**To ensure the test results of sample aliquots subject to various FTCs are directly comparable**, the FTCs were designed in such a way that the tests of all the aliquots of various FTCs were performed on the same run (RT-PCR) or same day (antigen) for the same sample. The exception is for cycle 0 in which a run-to-run normalization was performed.

### Rapid Antigen Test

**All samples were tested on the Hotgen Antigen test**, an LFA which detects the SARS-CoV-2 nucleocapsid antigen. The Antigen test was tested according to the instructions for use (IFU). Fifty microliters of specimen solution was carefully spiked onto the dry swab in the antigen kit then the swab was inserted into the extraction tube and mixed 15 times. After squeezing the swab against the tube wall 3 times, four drops were added to the device, and the result was read after 15 min. Photos of the Antigen test results were taken to compare the test line signal intensity throughout the cycles. Triplicates were tested for each condition and each sample.

**The positive antigen test signals** from the clinical remnant samples were used to generate a color intensity chart (Figure 2). Each signal was assigned a score from 0.5 (faint) to 10 (very strong), the higher the number the more SARS-CoV-2 in the sample. Negative was 0 (not shown in Fig. 2).

**Figure 2.**
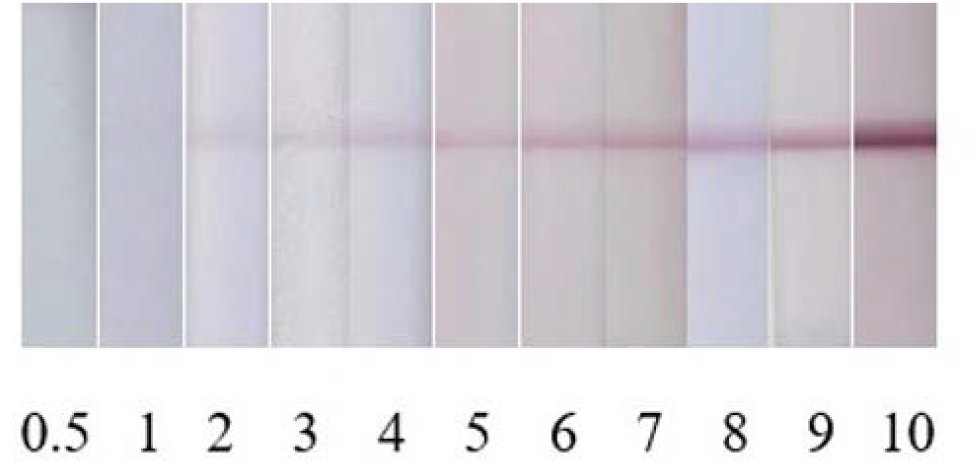
Color intensity chart of Hotgen Antigen test. The scale was made from enhanced Hotgen test image results and subjectively rated from weakest to strongest.

**All positive antigen test signals** in this study were compared to the color intensities in the chart and recorded from 0.5 (faint) to 10 (very strong).

### RT-PCR

**RT-PCR was performed using the laboratory-developed method**. Briefly, the samples were treated at 95°C for 5 minutes with 5 uL Proteinase K (BioLabs, Cambridge, MA) before RT-qPCR. The RT-PCR mix contains Reliance One-Step Multiplex RT-qPCR Supermix (BioRad, Hercules, CA), SARS-CoV-2-specific N gene primers (IDT, Coralville, IA), and human RNase P gene primers. RT-qPCR was run on the ABI 7500 Fast Dx Real-Time PCR (Thermo Fisher, Waltham, MA) instrument. Triplicates were tested for each condition and each sample. All data analysis was performed with Microsoft Excel for Microsoft 365 (Microsoft Corp. Redmond, Washington).

## Results

### FTCs Impact on Rapid Antigen Testing

**The Antigen test results of sample aliquots** subject to various FTCs are presented in Table 2 for clinical remnant samples and Table 3 for contrived viral culture fluid variant samples. The Antigen test results remained unchanged for most of the clinical remnant samples until 6 FTCs (Table 2). The positive signals decreased for two samples (FT07 and FT25) as soon as 2 FTCs, FT05 was weaker at 6 FTCs, two samples (FT04 and FT15) became weaker at 8 FTCs, most samples were slightly weaker by 10 FTCs with only 6 samples remaining the same intensity through all FTCs. For inactivated viral culture fluid samples, six of the nine listed in Table 1 were tested. All but two had no changes in antigen-positive signals throughout the FTCs (Table 3). In either sample type, all test results were positive on 100% of the positive samples, regardless of the number of FTCs, and the negative samples did not demonstrate any false positives.

**Table 2.** Antigen test results of clinical remnant samples after various freeze and thaw cycles.

| **Clinical**<br>**Remnant**<br>**Sample ID** | **0 FTCs** | **2 FTCs** | **4 FTCs** | **6 FTCs** | **8 FTCs** | **10 FTCs** |
| --- | --- | --- | --- | --- | --- | --- |
| **FT01** | 1 | 1 | 1 | 1 | 1 | 1 |
| **FT02** | 7 | 7 | 7 | 7 | 7 | 6.5↓ |
| **FT03** | 0.5 | 0.5 | 0.5 | 0.5 | 0.5 | 0.5 |
| **FT04** | 1.5 | 1.5 | 1.5 | 1.5 | 1↓ | 0.5↓ |
| **FT05** | 1.5 | 1.5 | 1.5 | 1↓ | 1 | 1 |
| **FT06** | 4 | 4 | 4 | 4 | 4 | 4 |
| **FT07** | 3.5 | 3↓ | 2.5↓ | 2.5 | 2.5 | 2↓ |
| **FT08** | 6.5 | 6.5 | 6.5 | 6.5 | 6.5 | 6↓ |
| **FT09** | 9 | 9 | 9 | 9 | 9 | 8.5↓ |
| **FT10** | 5.5 | 5.5 | 5.5 | 5.5 | 5.5 | 5↓ |
| **FT11** | 10 | 10 | 10 | 10 | 10 | 10 |
| **FT12** | 10.5 | 10.5 | 10.5 | 10.5 | 10.5 | 10.5 |
| **FT13** | 9.5 | 9.5 | 9.5 | 9.5 | 9.5 | 9.5 |
| **FT14** | 7.5 | 7.5 | 7.5 | 7.5 | 7.5 | 7↓ |
| **FT15** | 2 | 2 | 2 | 2 | 1.5↓ | 1↓ |
| **FT21** | 4.5 | 4.5 | 4.5 | 4.5 | 4.5 | 4↓ |
| **FT22** | 9 | 9 | 9 | 9 | 9 | 8.5↓ |
| **FT23** | 1 | 1 | 1 | 1 | 1 | 0.5↓ |
| **FT24** | 8.5 | 8.5 | 8.5 | 8.5 | 8.5 | 8.5 |
| **FT25** | 2.5 | 2↓ | 1.5↓ | 1.5 | 1↓ | 1 |

**Table 3.** Antigen test results of viral culture fluid samples after various freeze and thaw cycles.

| **Contrived Sample ID-**<br>**Concentration** | **0 FTCs\*** | **2 FTCs** | **4 FTCs** | **7 FTCs** | **11 FTCs** |
| --- | --- | --- | --- | --- | --- |
| **CS01**<br>1.73 x 10<sup>4</sup> TCID<sub>50</sub>/mL | 2 | 2 | 2 | 2 | 2 |
| **CS01**<br>3.45 x 10<sup>4</sup> TCID<sub>50</sub>/mL | 4 | 4 | 4 | 4 | 4 |
| **CS02**<br>1.73 x 10<sup>4</sup> TCID<sub>50</sub>/mL | 0.5 | 0.5 | 0.5 | 0.5 | 0.5 |
| **CS02**<br>3.45 x 10<sup>4</sup> TCID<sub>50</sub>/mL | 2 | 2 | 2 | 1.5↓ | 1.5 |
| **CS05**<br>4.53 x 10<sup>3</sup> TCID<sub>50</sub>/mL | 2 | 2 | 2 | 2 | 2 |
| **CS05**<br>9.06 x 10<sup>3</sup> TCID<sub>50</sub>/mL | 4 | 4 | 4 | 4 | 4 |
| **CS06**<br>2.85 x 10<sup>3</sup> TCID<sub>50</sub>/mL | 3 | 3 | 3 | 3 | 3 |
| **CS06**<br>5.7 x 10<sup>3</sup> TCID<sub>50</sub>/mL | 5 | 5 | 5 | 5 | 5 |
| **CS07**<br>5.75 x 10<sup>3</sup> TCID<sub>50</sub>/mL | 2 | 2 | 2 | 2 | 2 |
| **CS07**<br>1.15 x 10<sup>4</sup> TCID<sub>50</sub>/mL | 4 | 4 | 3.5↓ | 3.5 | 3.5 |
| **CS08**<br>2.25 x 10<sup>3</sup> TCID<sub>50</sub>/mL | 6 | 6 | 6 | 6 | 6 |
| **CS08**<br>4.5 x 10<sup>3</sup> TCID<sub>50</sub>/mL | 8 | 8 | 8 | 8 | 8 |

\*: The 0 cycle was defined as aliquots that were not subjected to FTCs in the study here, materials were already frozen when purchased.

### FTCs impact on RT-PCR Test of Clinical Remnant Samples

**As shown in Figure 3**, the Ct values of clinical remnant samples gradually increased with the increasing number of FTCs in all but one (FT02) of the positive samples (Figure 3A and 3B). The clinical remnant samples preserved in VTM had a mean slope of the linear regression line (FTC versus Ct) of 0.237 (Fig. 3A), whereas the mean slope for PBS samples was 0.405 (Fig. 4B), indicating a faster increase of Cts through FTCs for the PBS samples. When the viral load is low, the increase of Ct values in PBS samples becomes even more significant with increasing FTCs.

**Figure 3.**
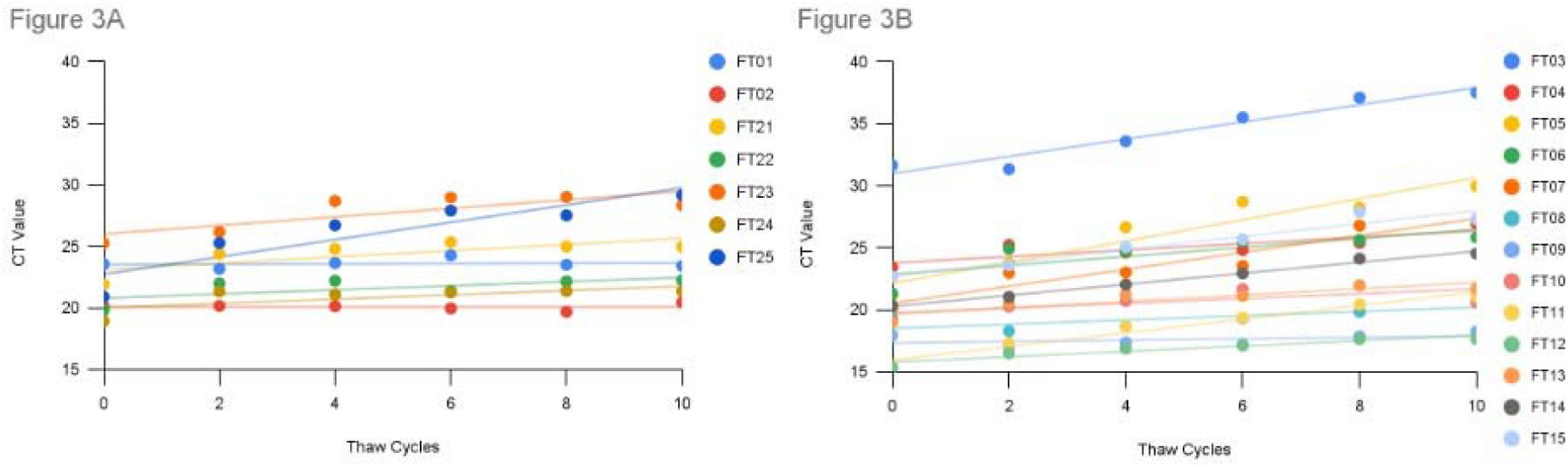
SARS-CoV-2 N-gene Ct values of clinical remnant sample aliquots after 2, 4, 6, 10 freeze and thaw cycles. A. Seven positive samples in VTM. B. 13 positive samples in PBS.

**Figure 4.**
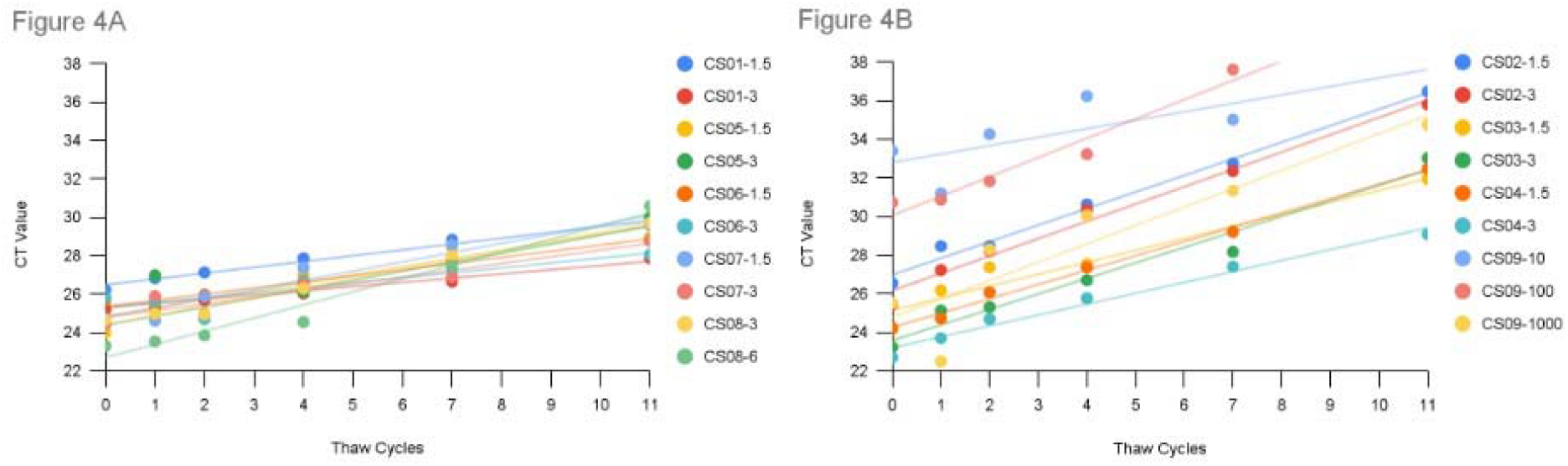
SARS-CoV-2 N-gene Ct values of viral culture fluid samples after 1, 2, 4, 7, and 11 freeze-thaw cycles. A. Five UV and Gamma-inactivated samples. B. Four heat-inactivated samples.

### FTCs impact on RT-PCR Test of Viral Culture Fluid Samples

**Linear regression was performed** on the average Ct values versus FTCs for the diluted viral culture fluid variant samples, where the slope represents the change in Ct value per freeze-thaw cycle (Figure 4A and 4B). The slopes steadily increased over time, reflecting the increasing degradation of the samples over the experiment time. Among 9 inactivated viral culture fluids variant samples, 5 were irradiation-inactivated (Figure 4A) and 4 were heat-inactivated (Figure 4B).

**The heat-inactivated contrived samples appear less resistant** to the FTC conditions (Fig. 4A) than the other irradiated-inactivated samples (Fig. 4B). As shown in Figures 4A and 4B, all other heat-inactivated samples had a much higher rate of Ct increase through FTCs. The Ct increase was evident even at the first few FTCs and with an average increase of 1.86 at 7 to 11 cycles by the end of the FTCs. Two samples (CS09-10 and CS09-100) became undetectable at cycle 11.

**The inactivated SARS-CoV-2 strains** were commercially purchased and are the main reference materials used in SAR-CoV-2-related research and product development. Their stability is thus very important. A standard good practice to keep the integrity of these reference materials is to aliquot them when the material is first thawed for use to avoid multiple freeze and thaw cycles. Our study showed here that even a few freeze-thaw cycles could have a negative impact on the integrity of SARS-CoV-2 nucleic acids, particularly the heat-inactivated ones. However, the samples studied here were diluted contrived samples. To find out if undiluted inactivated culture fluids would have better resistance to the FTCs, we performed FTC experiments on undiluted culture fluid samples. Aliquoted undiluted culture fluid samples were subjected to multiple FTCs (1, 2, 4, 7, and 11). After the FTCs, the samples were diluted to the concentrations used in the original dilution study (Table 3 and Figures 4A and B), and subject to RT-PCR analysis, no significant increase in Ct values was found even after 11 cycles for all the samples (Figure. 5).

**Figure 5.**
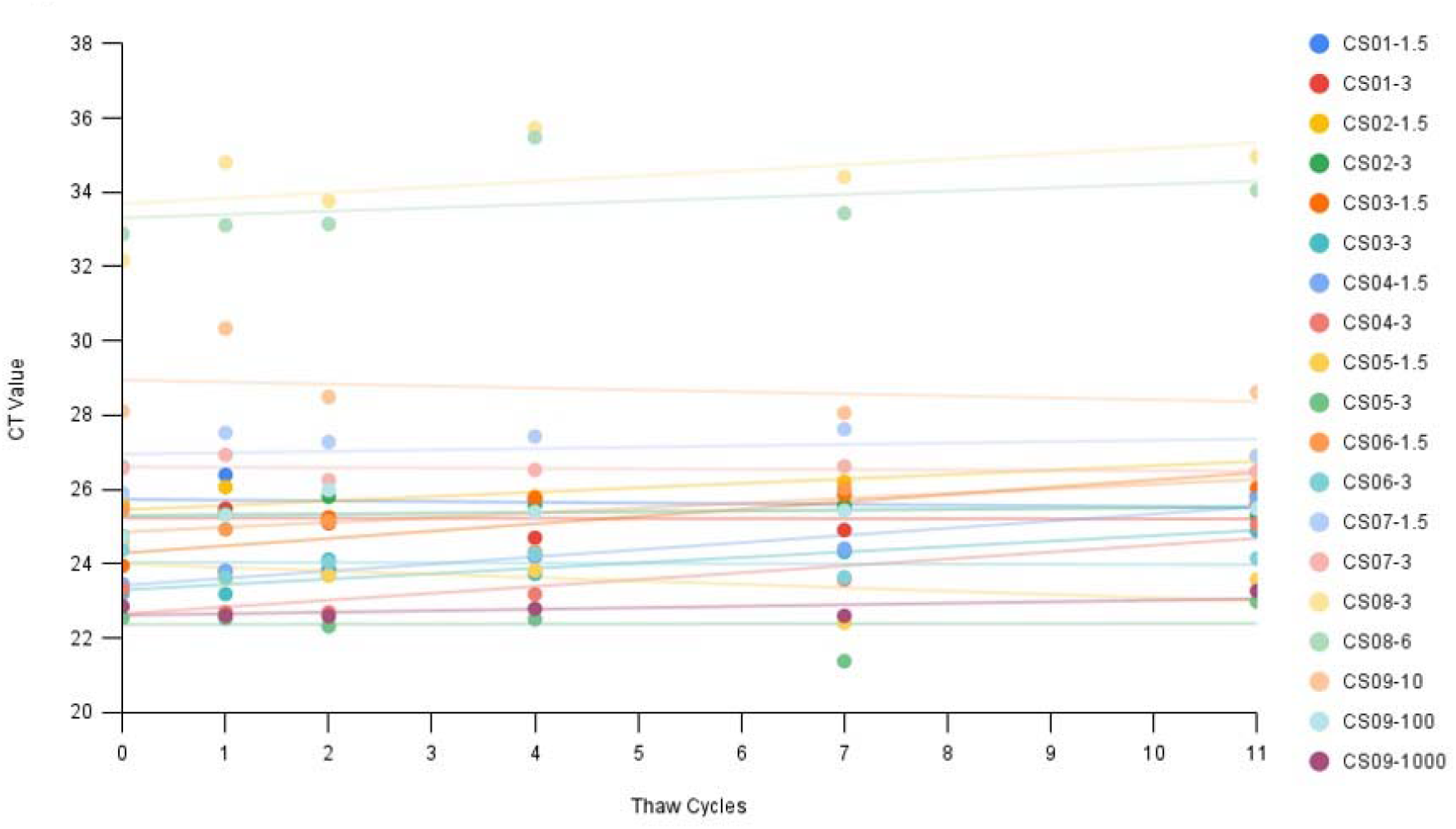
SARS-CoV-2 N-gene Ct values of viral culture fluid stock samples which underwent 1, 2, 4, 7, and 11 freeze and thaw cycles before being diluted for testing.

### Correlation between initial RT-PCR and rapid antigen test results

**Although both RT-PCR and rapid antigen tests are qualitative tests**, lower Ct values in RT-PCR and strong positive antigen test line signals usually indicate a strong viral load. In principle, these two test measures should correlate to each other at the active viral propagation phase when the same target gene/protein was used. In Figure 6, we compared the initial RT-PCR test results and antigen test results of 20 clinical remnant samples. As one can see, a correlation coefficient of -0.8839 is obtained, indicating a good negative correlation between the two test methods.

**Figure 6.**
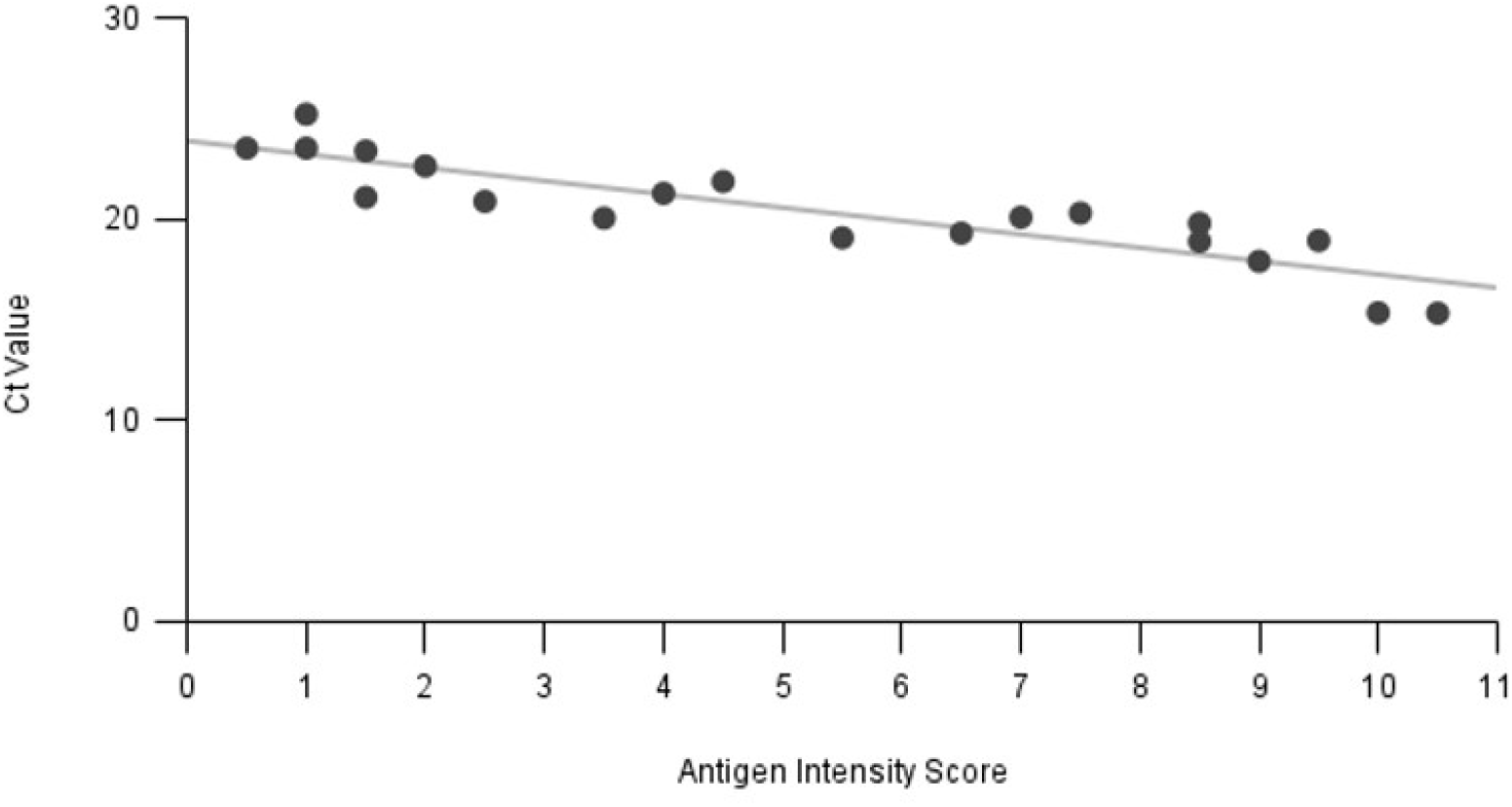
Correlation between Ct values and antigen test signal of 20 positive SARS-CoV-2 clinical remnants before FTCs.

## Discussion

**Ultra-low temperature has been a recommended shipping and long-term storage condition** for many human specimens including SARS-CoV-2 (https://www.cdc.gov/coronavirus/2019-nCoV/lab/guidelines-clinical-specimens.html). The study here as well as previous studies (1-5) demonstrated that the multiple FTCs can gradually damage the SARS-CoV-2 RNA, and thus should be avoided, if possible, particularly for the clinical specimens that were in PBS buffer and at low viral concentrations. The Ct values of RT-PCR could increase by more than 10, indicating severe nucleic acid degradation after multiple FTCs. In contrast, the SARS-CoV-2 antigen test is much less sensitive to the specimen subjected to FTCs. The damage on the antigen is undetectable before 6 FTCs.

**In this study, the RT-PCR test targets the nucleocapsid (N) gene**, whereas the rapid antigen test targets the N-protein. The use of the N-gene/protein in diagnostics has its merit. N-gene/protein is subject to fewer mutation events. Moreover, N-gene has much greater copies than other SARS-CoV-2 viral genes (12) and the copies of N-gene RNA and N-protein are closely correlated (13). The correlation was further confirmed with the clinical remnant samples tested here (Figure 6). The development of most SARS-CoV-2 rapid tests involves monoclonal antibodies (mAbs) that are peptide-specific to the epitopes of the SARS-CoV-2 N-protein. These antigen epitopes typically consist of 8-12 amino acids; a configuration that is less prone to alterations and, therefore, not extremely sensitive to conditions such as freezing and thawing. This may explain the stable Antigen test results on the samples experiencing many FTCs. In fact, antigen-antibody-based assay stability was also observed in several antibody tests (14-15). In contrast, the RNA is relatively less resistant to freeze and thaw cycles, especially at lower concentrations. The FTCs can break the human cells surrounding the virus leading to the release of RNAse. All the diluents used in the study for the viral culture fluids or the clinical remnant samples contain human cells. Moreover, the hydrolysis of the RNA phosphodiester backbone can also happen more evidently at a lower RNA concentration. The SARS-CoV-2 RNA vulnerability under various storage conditions may also be the cause of different test results reported from different labs, although the consensus appears the same that the multiple FTCs can eventually damage the SARS-CoV-2 RNA in clinical samples or samples contrived with human negative matrices. Interestingly, the viral culture fluid samples without any dilution can sustain multiple FTCs much better. High concentration and human cell-free conditions may provide a protection mechanism under multiple FTCs. Although it is always a good practice to aliquot the frozen reference materials at the first thaw and use, and avoid multiple FTCs during usage, it is good to know that multiple FTCs had minimal impact on the quality of the materials as long as they were stored frozen.

**Diluted heat-inactivated SARS-CoV-2 cultural fluid** was shown to be less stable than irradiation-inactivated ones after FTCs. Heat-inactivation can damage both SARS-CoV-2 RNA and proteins leading to diminished detection (11), whereas inactivation by irradiation tends to preserve the integrity of both antigen and nucleic acids better. Indeed, the study here showed that all heat-inactivated diluent samples had a much higher rate of Ct increase than the irradiation types after FTCs. This is very apparent in CS01 and CS02 which are of the same strain, both had the same LoD on the Antigen test the only difference between the two was the inactivation method. Both heat and irradiation are physical viral inactivation methods, heat inactivation primarily works by destroying the secondary protein structure (16), thus likely leading to more leakage of RNAse from human cells, RNAse would not be destroyed at the temperature the typical heat-inactivation protocol uses. On the other hand, irradiation inactivates the virus by breaking its nucleic acid chain. Irradiation damage on protein is minimal in general (17) but at a high intensity, it could also damage the RNAse to a certain extent (18). As long as the nucleic acid breakage induced by irradiation did not occur in the amplicon region, it would not affect the RT-PCR reaction. Therefore, irradiation-inactivated viral materials may provide a more robust option for certain research.

**An additional implication of this study is that the frozen samples** in an appropriate medium such as VTM or PBS used here could provide an alternative means for lateral flow antigen test developers to study the test performance when the resource of a clinical patient is scarce. The limitations of this study are that only one rapid antigen test was analyzed here. In addition, we also did not perform a systematic comparison of various types of VTMs, UTM, or other transportation mediums. Considering that multiple factors could affect both antigen and RT-PCR tests, we suggest that a similar study should be performed for any particular test system, should FTCs be concerned. The study can be performed at a much smaller scale, e.g., a smaller number of FTCs.

**Ultra-low temperatures have been a recommended shipping** and long-term storage condition for many human specimens including SARS-CoV-2 (https://www.cdc.gov/coronavirus/2019-nCoV/lab/guidelines-clinical-specimens.html). The study here as well as previous studies (2-6) demonstrated that the multiple FTCs can gradually damage the SARS-CoV-2 RNA, and thus should be avoided if possible.

**In conclusion, when making dilutions or testing clinical samples on rapid test kits**, freezing and thawing do not have a significant effect on the testing results. However multiple freezes and thaws should be kept to a minimum as RT-PCR testing reveals it can cause significant degradation of RNA, especially at low concentrations, and possibly some degradation to antigens long-term.

## Data Availability

All data produced in the present work are contained in the manuscript

## Acknowledgments

**All authors declare** that they have no conflicts of interest.

**This study was funded** by XYZ Laboratory which is self-funded.

**We would like to thank** Beijing Hotgen Biotech Co. Ltd. for providing the lateral flow rapid antigen test (Antigentest) for this study. Thanks Dr. Wei Zeng, Ruifeng Xiao, and Jun Zhang for critical reading of the manuscript. WZ and KG for contriving the ideas, planning the experiment, and reviewing the manuscript, HNH for writing the manuscript, making the figures and tables, performing the variant FTC and antigen experiments; HW for assisting with the initial draft of the manuscript, performing the clinical remnant FTC, and antigen experiments; DZ for initial data analysis; DJ, BL, HX, and CZ for performing the PCR experiments.

